# Dementia Training for Healthcare Professionals: A Systematic Policy and Evidence Review

**DOI:** 10.1101/2025.01.15.25320582

**Authors:** Sedigheh Zabihi, Saskia Delray, Malvika Muralidhar, Sube Banerjee, Clarissa Giebel, Karen Harrison Dening, Yvonne Birks, Rachael Hunter, Mohammed Akhlak Rauf, Charlotte Kenten, Madeleine Walpert, Claudia Cooper

## Abstract

**Objective:** To review policies and research on the effectiveness of healthcare professionals’ dementia training

**Design:** Policy and systematic review

**Setting:** Healthcare services

**Participants:** Healthcare professionals

**Intervention:** Training

**Measurements:** We reviewed relevant English policies and searched electronic databases for primary research studies (between 2015-2024) evaluating dementia training for healthcare professionals. We assessed risk of bias using the Mixed Methods Appraisal Tool, prioritising studies scoring 4+, of interventions supported by Randomised Controlled Trial evidence, and reported outcomes using Kirkpatrick’s framework. We consulted professional stakeholders in a focus group regarding how findings might inform practice.

**Results:** Sixteen policies and related documents highlighted concerns about the limited implementation of the Dementia Core Skills Education and Training Framework (DCSETF). We reviewed 63 primary research studies. Only one study met priority criteria; it evaluated a Train-the-Trainer (TTT), team-based reflective practice model, which improved primary care nurses’ and doctors’ learning, and self-reported practice over ≥3 months. Higher quality, controlled studies evaluated a TTT programme for hospital staff, improving client outcomes (agitation) over ≤5 days; an expert-led two-day interactive training for inpatient nurses that reduced role strain; and an expert-led, nine-week, occupational therapy-derived training programme that improved retirement community staff strategies for client activity engagement. Eight focus group attendees considered time a limiting factor to evidence implementation, but valued group training to share experiences; and TTT models to enable tailoring to local contexts.

**Conclusions:** By increasing reach of dementia training and embedding learning in practice, Train-the-Trainer models could increase care quality and DCSETF implementation.

## Introduction

Around one million people have dementia in the UK, and this is expected to rise to 1.4 million by 2040 [1]. Almost all primary, and most secondary care healthcare workers, provide care to people with dementia. This requires specific skills and knowledge, and inadequate training and support can lead to negative outcomes for workers, people with dementia and their families [2].

The Dementia Core Skills Education and Training Framework (DCSETF), first published in 2015, outlines learning outcomes that health and social care workers in England are expected to accomplish [3]. A review of international standards cited these as among the strongest national guidelines underpinning dementia workforce training development. However, subsequent service audits in England have indicated that these standards are often unmet [4,5], in part due to lack of access to relevant training [6].

Developing an appropriately skilled care workforce is a national policy priority [7]. Submissions to the Darzi report into the state of the NHS highlighted concerning variations in post-diagnostic care quality, and evidence-based treatment availability, which improved workforce training could address [8].

To inform future policy regarding dementia training for healthcare professionals, we updated a previous systematic review [9], by exploring the evidence base for dementia training and how policy has evolved since publication of the DCSETF. Our Research Questions (RQ) were:

1. What policies (since 2015) have relevance to dementia training for healthcare professionals?
2. What is the current evidence on how dementia training is best delivered to healthcare workers to improve dementia care and worker wellbeing?
3. How do stakeholders consider current evidence might inform future policies and practice to positively change healthcare workforce training?

## Methods

### 2.1 Defining social and policy context

The study group agreed sources (Table S1) and terms to search English policy and grey literature (from 2015) relevant to dementia skills training for healthcare professionals to respond to RQ1. Search terms were: “health” and “training” and “dementia”. SZ and CC independently identified potentially relevant documents in November 2024, then discussed which to include based on relevance to the research question. Findings were discussed with the wider study group to develop descriptions of policy contexts and challenges as our empirical and conceptual understanding evolved during the review.

### 2.2 Systematic review

The study was conducted in line with Preferred Reporting Items for Systematic Reviews and Meta-Analyses (PRISMA) recommendations [10] and registered in PROSPERO (CRD42024509026).

#### 2.2.1 Search Strategy

Databases including PubMed, Embase, Scopus, CINAHL and the Cochrane library were systematically searched from 01 December 2015 (to update the previous review [9]) to 20 February 2024. For the database search, terms related to education OR training were combined with staff AND dementia (Table S2). Searches were restricted to studies published in English.

#### 2.2.2 Eligibility Criteria and Study Selection

After excluding irrelevant titles and abstracts, two reviewers independently screened full-text papers against eligibility criteria to identify studies for inclusion using Covidence software. Discrepancies were resolved through discussion. We included primary research studies reporting the effectiveness of any training or educational intervention for healthcare workers, focused on development of dementia-specific knowledge, values and skills to improve dementia care. We excluded dissertations, conference and meeting abstracts. We excluded studies that trained staff to deliver specific, manualised interventions, or where training focused on the delivery of care for conditions comorbid with dementia, or delivery of end-of-life care.

#### 2.2.3 Data Extraction and Quality Appraisal

A standardised form was developed to extract data from included studies describing: population characteristics; training intervention; control conditions (if applicable); and outcomes. We used the Mixed-Methods Appraisal Tool (MMAT) to assess study quality [11]. Each study was independently assessed by two authors and disagreements were resolved by a third author. Studies were scored out of five and studies attaining four or more were considered high-quality [12].

#### 2.2.4 Data Analysis

We categorised studies by intervention settings and delivery models. Within each category, we narratively synthesised findings, mapping reported outcomes to Kirkpatrick’s framework: learner’s reaction to and satisfaction (Level 1); knowledge, skills, confidence, and attitude change indicating learning has occurred (Level 2); staff behaviour or practices (Level 3); and patient-related outcomes (Level 4) [13]. We report between group differences for controlled studies, within group changes for single group studies, and all relevant outcomes.

### 2.3 Stakeholder meeting

We held one online workshop on November 18^th^ 2024, to which we invited healthcare professionals from diverse roles working with people with dementia, identified via DeNPRU-QM national networks. To respond to RQ3, we presented findings emerging from our evidence review and asked the group to consider how feasible and useful the interventions for which we identified high-quality evidence might be in practice. We outlined findings from the policy review and invited the group to identify further relevant sources or documents. Verbatim notes were circulated to attendees after the group, inviting corrections and clarifications.

## Results

### 3.1 Defining policy context

We identified sixteen relevant policies (Table S3), with no further sources provided by the stakeholder group. These described an expectation that healthcare workers will receive role-appropriate dementia training [14]. The DCSETF defines training objectives for all healthcare workers (Tier 1), those working regularly with people with dementia (Tier 2) and care leaders (Tier 3). NHS England publications discussed “Training Well” as a component of the Dementia Well pathway [15], and culturally competent care [16].

Two reports advocate Tier 1 training for all hospital staff, wider roll out of Tier 2 and 3 training, and organisational annual reporting of staff training levels [17,18]. A 2018 National Collaborating Centre for Mental Health report recommends that commissioners agree training plans and associated costs with providers, and ensure sufficient workers are trained to appropriate levels [19]. A 2023 Royal College of Psychiatrists audit suggested implementation of these recommendations was patchy [5]. Reports have highlighted training needs around comorbidities [19], behaviours indicating distress, reducing the need for antipsychotics, promoting freedom of movement, and holding challenging conversations [20]. The Royal College of Nursing [21] recommended that training should be team-based, so skills are shared; and include lived experience content.

### 3.2 Systematic review

We included 63 studies meeting the eligibility criteria (Figure S1).

#### 3.2.1 Study characteristics

Twenty-six (42%) included studies were rated as high-quality (4+ on MMAT). These are described in Table 1. Only one study met our *a priori* criteria for prioritisation (in the primary care studies section). We included non-randomised quantitative (n=12) and mixed-method studies (n=11), randomised controlled trials (RCT, n=2) and one qualitative study. Details of studies scoring <4 on MMAT are in Table S4; and briefly summarised below.

**Table 1.**
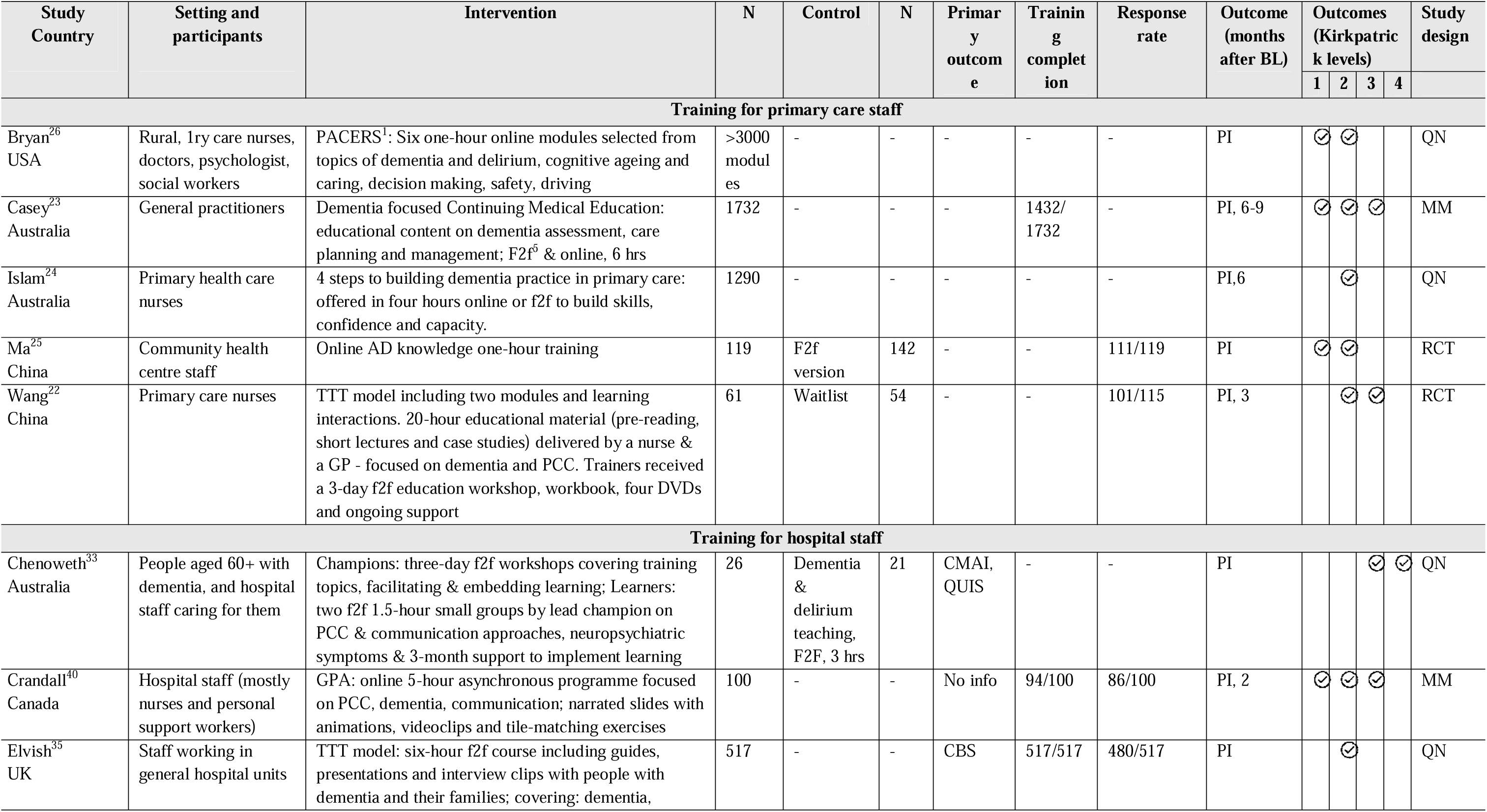
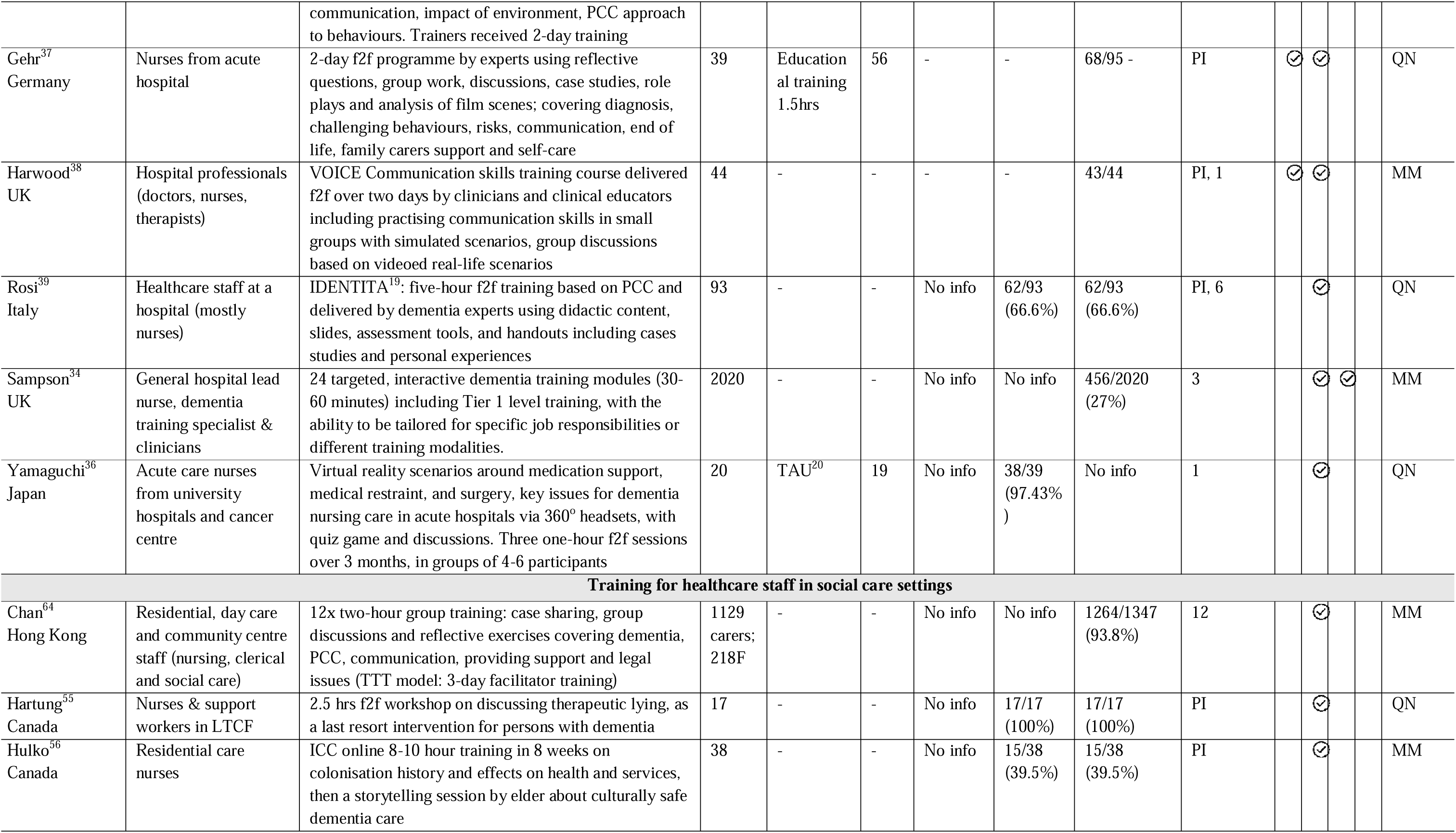
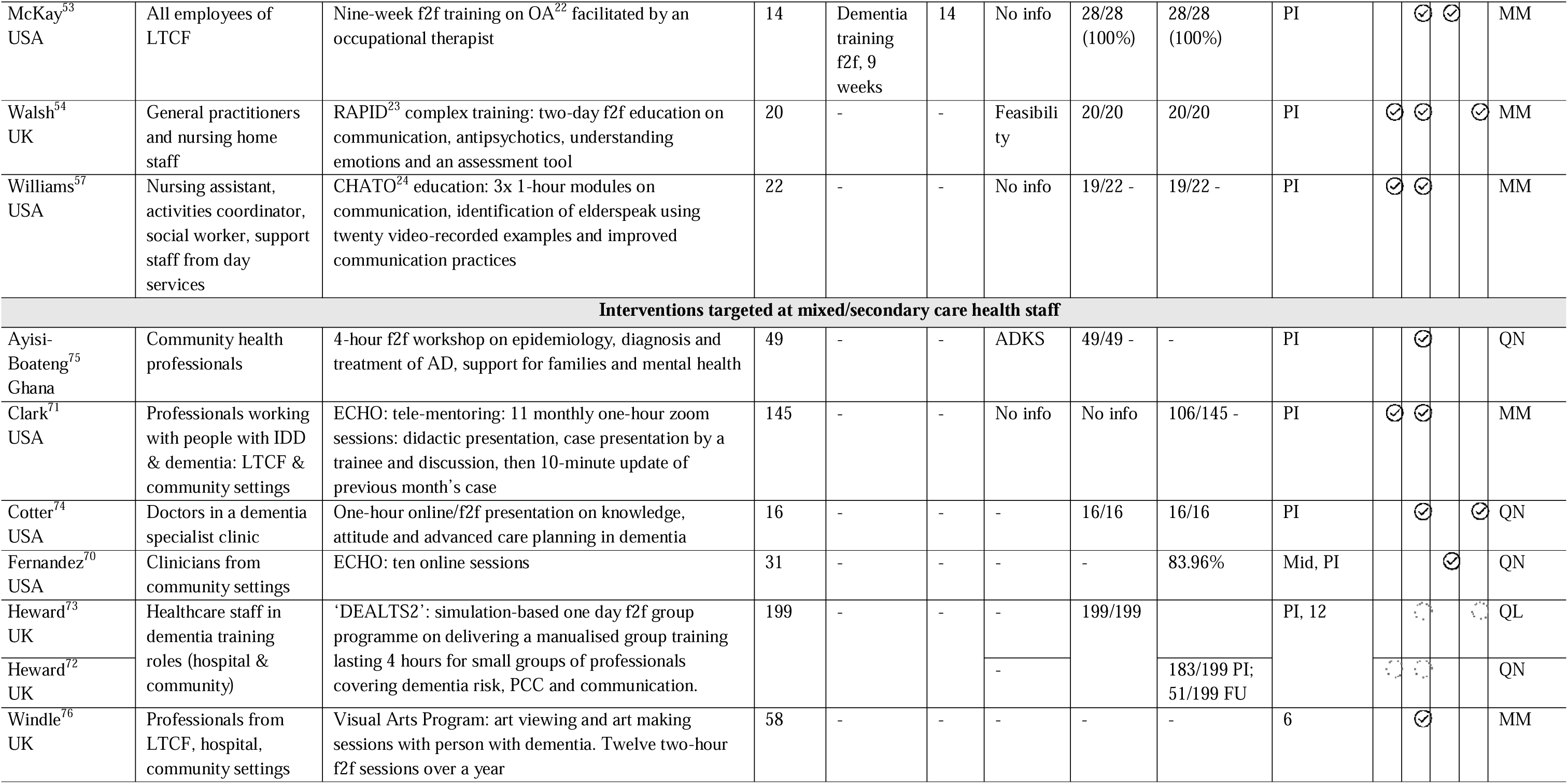
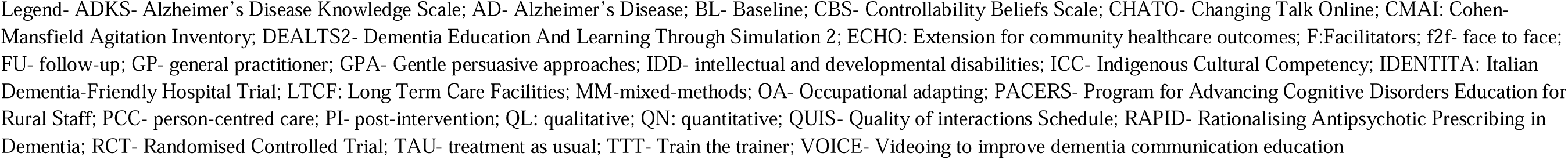
Characteristics of the studies rated as higher quality.

We divided included studies into four groups, based on intervention settings: (1) primary care; (2) hospital; (3) social care settings (residential, nursing home or day care services) and (4) secondary care or staff groups across primary and secondary care. We further grouped studies by intervention delivery model: Train-the-Trainer (TTT), expert-delivery (clinical or lived experience expert), tele-mentoring, arts-based, self-directed, asynchronous, in-person or online lecture-based (didactic). Figure 1 summarises evidence of effectiveness from higher-quality controlled studies.

**Figure 1:**
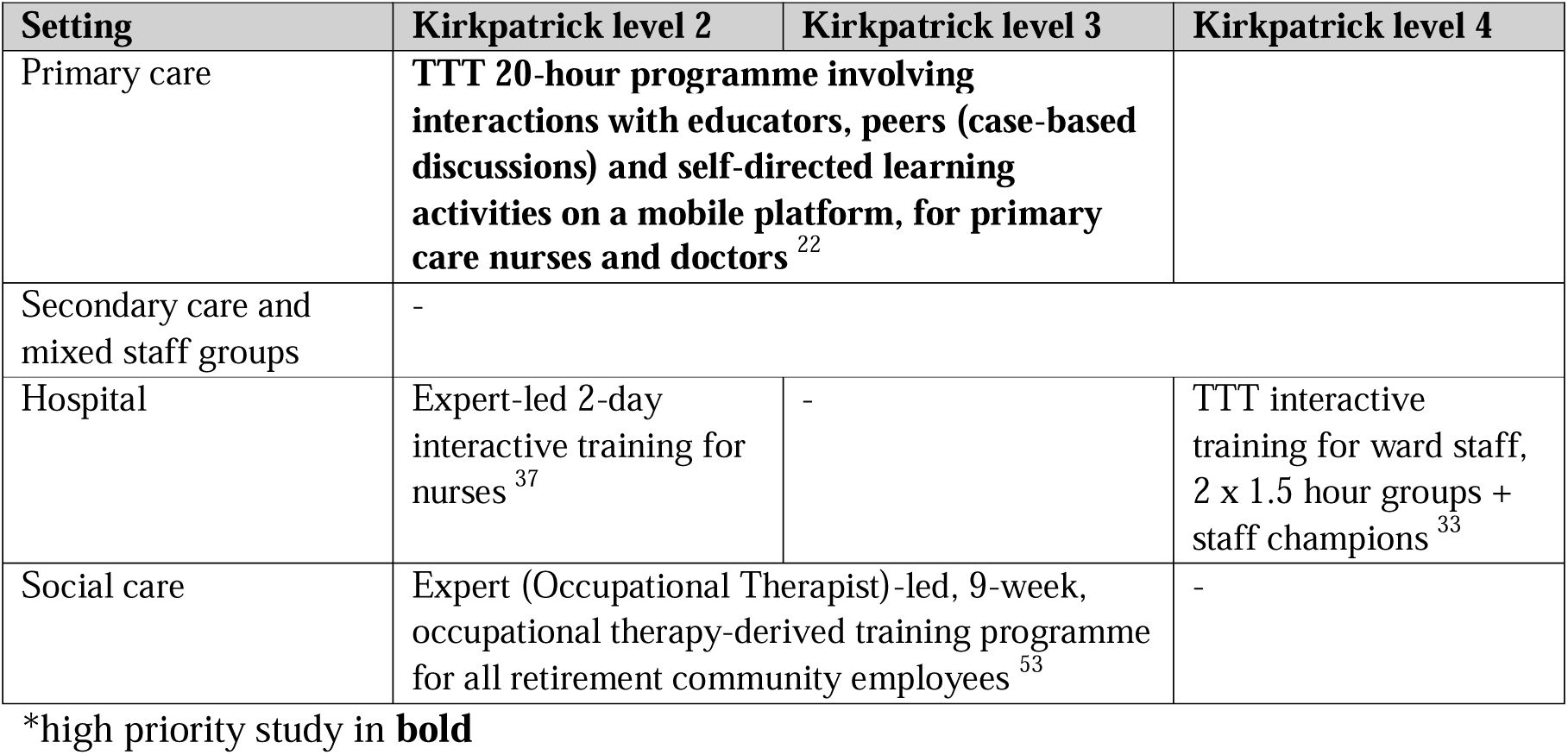
Summary of interventions for which significant between group differences were reported, by site and Kirkpatrick evidence levels 2-4, from high quality, controlled studies*

#### 3.2.2 Training for primary care staff

##### High-quality studies (n=5)

###### Train-the-Trainer models

The only study that met our criteria for higher priority evidence, an RCT, evaluated a web-based training, comprising two modules and learning interactions with educators, peers and self-directed learning activities, delivered in a TTT model. Compared to a waitlist control group, nurses and general practitioners (GPs) allocated to the intervention training improved on scores of knowledge (Alzheimer’s disease knowledge scale [ADKS]; F= 31.35; P<0.001), attitudes (Dementia Attitudes Scale [DAS]; F=20.57; p<0.001) and the intent to change practice towards early dementia diagnosis (self-developed scale; P<0.05) up to three months later. In focus groups, participants described a positive impact on their dementia care practice, including screening for cognitive impairment and referring to memory services more frequently (level 2 and 3) [22].

### Asynchronous or lecture-based models

An Australian study evaluated a national, dementia-focused Continuing Medical Education (CME) programme for GPs. It comprised at least six hours of structured educational content, with two-thirds interactive or experiential content (case studies and discussion). It was offered as online (six one-hour modules), large, or small group face-to-face workshops (four 90-minute sessions). Participants’ knowledge (mean difference= 2.2; p<0.001), confidence (mean difference= 2.1; p<0.001), awareness of dementia (mean difference= 0.9; p<0.001) and self-reported practice (e.g. referring to support services and using non-pharmacological strategies) (mean difference= 1.3; p<0.001) improved post-intervention (level 2 and 3) [23]. A second Australian training programme for primary care nurses aimed to promote evidence-based approaches to dementia detection, diagnosis and support. It was available in online (four one-hour asynchronous modules) and in-person (one 3.5-hour group session) modalities. In 1,290 primary care nurses who participated, training improved knowledge, confidence and perceived importance of dementia diagnosis (bespoke measure; p<0.001) (level 2). Improvement after in-person training were greater, relative to online delivery over six months [24]. Finally, an RCT, which reported Level 2 evidence, compared online and in-person versions of a one-hour lecture on Alzheimer’s disease, for community care staff. There were no significant differences in ADKS scores between groups post-intervention (p>0.05) [25].

### Self-directed asynchronous models

Program for Advancing Cognitive Disorders Education for Rural Staff (PACERS) was a USA programme for healthcare professionals working in rural areas, developed in six one-hour online asynchronous modules. Participating staff reported self-perceived ability to apply learnt knowledge and skills (mean>4 /5) (level 2) [26].

#### Lower quality studies (n=7, Table S3)

One lower quality study reported that after a four-hour in-person training on Advance Care Planning (ACP), primary care clinicians documented more ACPs (level 4) [27]. An online course on interprofessional communication for primary care professionals [28], face-to-face TTT model of Person-Centred Care (PCC) training for primary care nurses [29], online course on medication and behaviour management with ongoing support from geriatric experts for primary care nurses [30] and a hybrid postgraduate dementia course for primary care staff ^31^ positively impacted care. The postgraduate hybrid dementia [31], online course on interprofessional communication [28], face-to-face TTT model of PCC training [29] and another online tool to assist with diagnosis and care [32], improved knowledge, attitudes and confidence (level 2) in primary care staff.

#### Summary of higher quality evidence

- A high-priority study evaluated a 20-hour, **TTT programme** involving interactions with educators, peers (case-based discussions) and self-directed learning activities on a mobile platform, for primary care nurses and doctors; the intervention improved self-reported change in dementia care practice (level 3) and knowledge and attitudes to dementia (level 2) for up to three months, relative to the control condition.
- In Australian single group studies of interventions provided in a choice of **online asynchronous modules or face to face formats**, a national training programme for GPs (interactive sessions, including case-based discussions) improved knowledge and self-reported practice (levels 2 and 3) for up to nine months; and a programme for nurses improved knowledge for up to six months (level 2).

#### 3.2.3 Training for hospital staff

##### High-quality evidence (n=8)

###### Train-the-Trainer models (n=3)

Of the three studies evaluating TTT models, only Chenoweth [33] included a control condition, in a pilot study, testing a PCC training programme. Selected staff were trained to be ‘champions’ or trainers, in three-day workshops. A champion and study investigator then facilitated two 1.5-hour small group sessions for nurses and allied health staff; and supported implementation of learning over three months. In short hospital stays (4-5 days), in patients cared for by trained staff, staff-rated agitation measured by Cohen-Mansfield Agitation Inventory (incidence rate ratio [IRR] = 0.84, 95%: 0.70 to 0.99; p= 0.04), delirium (odds ratio [OR]= 0.1, 95% CI: 0.005 to 0.85); p= 0.033) and iatrogenic harms (p=0.02) decreased (level 4) and quality of care measured by Quality of Interactions Schedule (QUIS; IRR= 1.38, 95% CI: 1.06 to 1.80; p= 0.017) increased (level 3), compared to the control condition. Only improvements on iatrogenic harms were maintained for 8 days [33].

In a second study, Sampson [34] evaluated a structured training package, delivered by ward staff after training. It comprised 24 interactive dementia modules (30-60 minutes) including DCSETF Tier 1 and 2 topics. Modules were standardised but could be tailored to specific roles and delivered in-person, groups or one-to-one. After training, Sense of Competence in Dementia care [SCID] scale scores improved (mean score 43.2 to 50.7; p<0.001) (level 2), and observations indicated improved quality and quantity of person-focused interactions on wards where staff received training (level 3), over three months [34]. The final TTT study evaluated a six-hour course delivered by trained staff facilitators, which included videoclips of people affected by dementia. After training, participants scores improved on the Knowledge in Dementia scale (KIDE) (r=-0.8; p<0.001), Confidence in Dementia [CODE] (r=-0.96; p<0.001) and self-efficacy in managing challenging behaviours (Controllability Beliefs Scale; r=0.51; p<0.001) (level 2) [35].

### Expert-delivery models (n=4)

These studies evaluated expert-led, in-person group interventions that sought to change knowledge, confidence and attitudes through exposure to lived experience, simulations, and role-play activities in training programmes lasting from three hours to two days. All aimed to improve communication and PCC skills, and management of challenging behaviours, in the hospital environment. All reported evidence at Kirkpatrick Level 2. Two studies included a control condition. These included a Japanese study evaluating the impact of three, one-hour sessions for acute care nurses, including virtual reality simulations of key issues for dementia nursing care, followed by games and discussions. There were no significant between-group difference in outcomes (Dementia Nursing Competency Scale in Acute Hospitals [DNCS- AH]; Staff Experiences of Working with Demented Residents questionnaire [SEWDR]) KIDE; and DAS) up to one month after training [36].

Two studies evaluated two-day training programmes facilitated by clinical experts in Germany and the UK which improved confidence of attendees in dementia care. The German programme for nurses, evaluated in a controlled study, was facilitated by nursing and gerontology experts using reflective questions, facilitated discussions, case studies, role-plays and film scenes analysis. Compared with the control condition, perceived strain (Modified Nursing Care Assessment Scale [M-NCAS]; F=7.70; p=0.007) and confidence (CODE; F=18.05; p<0.001) improved in the training group, while attitudes did not (M-NCAS; F=1.87; p=0.17) [37]. The UK programme, VideOing to improve dementia communication education (VOICE) was co-produced with stakeholders using similar learning strategies.

Participants improved in confidence (CODE; mean difference=5.5; 95% CI: 4.1 to 6.9; p<0.001) and knowledge of communication in dementia (unvalidated measure; 1.5; 95% CI: 1 to 2; p<0.001) [38]. A briefer, five-hour programme in Italy that used didactic teaching, and included learning from cases studies and personal experiences was associated with improved knowledge (Dementia Knowledge–20 [DK-20]; F=41.39; p<0.001), confidence (CODE; F=8.64; p<0.001), and increased personhood subscale scores on the approaches to dementia questionnaire (ADQ; F=3.85; p=0.02) up to 6 months after training [39].

### Self-directed asynchronous models (n=1)

Crandall evaluated 2-3 hours of online, self-directed PCC training over six weeks comprising narrated slides, videoclips and exercises in a mixed-methods, single group study [40]. In focus groups, participants reported using learnt, more tailored PCC for challenging behaviours. Knowledge (bespoke measure; F=14.78; p<0.001), self-efficacy (Self-Perceived Behavioural Management Self-Efficacy Profile [SBMSEP]; F= 34.53; p<0.001) and competence in dementia care (Sense of Confidence in Dementia Care Staff scale [SCIDS]; F=61.88; p<0.001) increased post-intervention; improvements were maintained at 6 weeks (level 2) [40].

### Lower quality studies (n=14, Table S4)

These findings aligned with higher quality studies reported above. They reported: level 4 evidence for an intervention described above [39]: clients of trained professionals maintained their functional ability after discharge [41]. They reported level 3 evidence that staff trained in PCC approaches used more person-centred communication [42,43]; and level 2 evidence for interventions that improved staff knowledge, attitudes and confidence [42–52].

#### Summary of higher quality evidence

- Interactive training packages delivered by ward staff using **TTT models** improved outcomes for patients with dementia (level 4) in a controlled study, and in staff practice (level 3) and learning (level 2) in a single group study.
- In controlled studies evaluating **expert-delivered, in-person group interventions**, three, one-hour groups for acute care nurses in Japan to address critical issues in dementia care did not change level 2 outcomes; while a two-day workshop involving interactive, case-based methods did reduce strain and improved confidence, though attitudes did not change post-intervention (level 2). Two single group, higher quality interventions lasting 5-6 hours improved level 2 outcomes.
- A single group study found that 2-3 hours of interactive online training was associated with improvements in staff practice (level 3) and learning (level 2) for up to six weeks.

#### 3.2.4 Training for healthcare staff in social care settings

These studies outline dementia training that included staff from healthcare disciplines working in social care including residential, nursing homes and day centre services.

### High-quality studies (n=6)

#### Clinical expert delivery models (n=3)

These small studies evaluated interventions delivered by occupational therapists, academic nurses and clinical pharmacists. McKay et al. [53], in a controlled study, tested a nine-week, occupational therapy-facilitated training programme, teaching continuing care retirement community staff (nurses, social care and other professionals) strategies to engage clients in meaningful activities. Compared to the active control condition, those allocated to the intervention improved in relative mastery (Relative Mastery Measurement Scale [RMMS]; p<0.05) and team development scores (Team Development Measure [TDM]; p<0.05) (level 2). Participants reported providing more PCC in qualitative interviews (level 3) [53].

Researchers in Ireland developed the Rationalising Antipsychotic Prescribing in Dementia (RAPID) intervention for nursing home staff and GPs, to reduce inappropriate antipsychotic prescribing. GPs received training on assessment and treatment of behavioural and psychological symptoms of dementia, and an antipsychotic deprescribing algorithm from a pharmacist. Three months after the programme, prevalence of antipsychotic receipt by residents decreased without substitution for other drugs or worse clinical outcomes (44% to 36%; not statistically compared due to small sample). Four GP participants reported that the programme was useful and well-suited to their busy schedules [54].

Finally, a Canadian study evaluated a 2.5-hour workshop delivered by academic nurses for nurses and support workers in hospitals and long-term care facilities on therapeutic lying in the context of PCC. Participants’ sense of competence (SCIDS; p=0.24), or attitude towards lying to people with dementia (bespoke measure; p=0.44) did not change post-intervention [55].

### Lived experience expert delivery model (n=1)

A Canadian study evaluated an eight-hour, asynchronous online training then a storytelling session and talking circle with elders from indigenous communities on dementia and nursing care. Fifteen nurses who received training applied more culturally-safe care plans (measured using a vignette) (level 3), and reported increased hope (ADQ; mean score changed from 28.27 to 31.73), and self-perceived knowledge of cultural safety (p<0.01) [56].

### Self-directed asynchronous model (n=1)

Williams [57] evaluated Changing Talk Online (CHATO) in adult day services. The online training included three one-hour modules on effective communication, completed over four weeks. Modules asked participants to identify instances of “elderspeak” in videoclips as part of the training. Nineteen participating adult day service staff improved in knowledge (self-perceived measure; mean difference=15; p<0.001), confidence (CODE; mean difference=1.9; p=0.037) and recognition of elderspeak communication (self-perceived measure; p=0.025).

### Train-the-Trainer model (n=1)

In a single group evaluation in Hong Kong, residential and community care staff were trained to deliver PCC-focused training. The training included 12, two-hour training sessions for groups of around six learners over six months using case sharing, group discussions and reflective exercises. After twelve months, participants’ scores on knowledge (Dementia Knowledge Assessment Scale [DKAS]; t=20.4; p<0.001), attitudes towards dementia (DAS; t=26.6; p<0.001), sense of competence (SCIDS; t=21.4; p<0.001) and job satisfaction (Satisfaction with Nursing Care and Work Assessment Scale [SNCW]; t for facilitators=4.8; t for learners=14.6; p<0.001) increased in facilitators and learners (level 2) [58].

### Lower quality studies (n=13, Table S3)

Lower quality study evidence aligned with higher quality evidence. A self-directed online 5.5-hour course on psychosocial interventions improved neuropsychiatric symptoms in people with dementia (level 4) [59]. We identified level 2 and 3 evidence for a two-year, monthly interactive PCC training [60], an online five-hour course on management of challenging behaviours [61], and twelve-session online PCC courses delivered by dementia experts [62,63]. In these studies, healthcare staff from different background implemented suggested behavioural approaches [60–63]. We also found level 2 evidence which showed they improved knowledge, attitudes and confidence in care for: a twelve-hour online TTT training [64], a one-hour in-person course covering communication strategies by a speech pathologist [65], an online six-session course on communications and managing emotions [66], ECHO [67], and VOICE [68].

#### Summary of higher quality evidence

- One small, controlled study reported that a nine-week, occupational therapy-derived training programme, facilitated by an occupational therapist improved level 3 and 2 outcomes.
- In small, single group studies, immediately after intervention delivery: a pharmacist-led intervention reduced GP antipsychotic prescribing; asynchronous online learning combined with a storytelling group led by indigenous elders increased nurses use of culturally safe care plans, hope and knowledge about cultural safety; and an interactive self-directed learning intervention reduced elderspeak in daycare service staff.
- A TTT programme enhanced staff knowledge and confidence up to 12 months after delivery of a training package comprising 12, two-hour small group sessions over six months to residential and community care staff in a single group study.

#### 3.2.5 Interventions for mixed/ secondary care health staff

##### High-quality studies (n=6)

###### Tele-mentoring models (n=2)

Two studies evaluated *Extension for community healthcare outcomes (ECHO),* a tele-mentoring model previously demonstrated to improve staff and patient-related outcomes in an RCT [69]. Fernandez et al. [70] evaluated an ECHO programme for community clinicians (mainly family medicine residents), comprising eleven monthly one-hour zoom sessions delivered by an expert interdisciplinary panel (geriatrician, geriatric psychiatrist, nurse practitioner with geriatric expertise, geriatric pharmacists, and a social worker) to medicine and psychiatry residents (doctors) and physician assistants. Each session involved a 20- minute didactic presentation, 30-minute clinical case presentation, group discussion, then a 10-minute update on the previous month’s case. The proportion of clinicians intending to change their prescriptions based on the sessions meetings increased during the intervention period (level 3) (self-perceived measure; p[=[0.04) [70]. Clark et al. [71] adapted ECHO for health and social care professionals working with older adults with Intellectual and developmental disabilities (IDD) in residential, primary care, community and academic settings. They evaluated ten one-hour sessions of didactic and case study discussions on IDD and dementia over twelve months. In 145 primary and community professionals recruited from over 20 organisations, knowledge of dementia improved (self-perceived measure; beta= 0.69, 95% CI: 0.23 to1.15) after training delivered by the Hub team (partnership between academic institutions, community organizations, and service agencies) (level 2) [71].

### TTT model (n=1)

Dementia Education And Learning Through Simulation 2 (DEALTS2), is a simulation toolkit commissioned by Health Education England (HEE). It comprises a fully structured course, deliverable in a single, four-hour session to small staff groups, covering dementia risk reduction, PCC, scenarios for experiential learning; and a one-day TTT course. Heward et al. [72] delivered the DEALTS2 TTT course to 199 dementia trainers in hospital and community settings. In qualitative interviews, trainers noted improvements in clinical practice and empathy in trained staff [73] up to nine months after the TTT course. Participants’ knowledge (self-perceived 5-point Likert scale) improved after training (p<0.001) in all areas (dementia risk factors, lifestyle changes to reduce risk, person-centred approaches, communication and interaction, humanised approaches, and signposting to sources of support) with most maintained for up to a year. Improvements in confidence was also maintained over a year [72].

### In-person or online lecture-based (didactic) models (n=2)

Cotter et al. [74] developed a one-hour programme to train healthcare professionals in ACP, online or in-person. It increased staff knowledge (self-perceived measure; p=0.002), and confidence discussing ACP (developed measure; p=0.03), and belief it can improve outcomes (developed measure; p=0.03) immediately after the presentation. Documentation of advance directives (13.6% to 19.7%; p= 0.04) and medical orders for life-sustaining treatments increased (11.0% to 19.0%; p= 0.006) [74]. Secondly, a one-day large group, didactic workshop for healthcare professionals in Ghana, on Alzheimer’s disease symptoms and management was associated with increased scores of participants on the ADKS (mean [SD]_pre_= 19.8 [4.3]; mean [SD]_post_= 23.2 [4]) [75].

### Arts-based intervention models

A UK, visual arts programme for family and professional carers from care homes and dementia care services included twelve, two-hour weekly sessions of art viewing and making with people with dementia. Qualitative interviews with professional carers showed that the programme helped them see the capabilities of the person with dementia, but quantitative outcomes did not reflect this (ADQ; p= 0.99) [76].

### Lower quality studies (n=2)

One study evaluated a series of three in-person two-hour workshops on clinical updates in dementia-related behaviours and principles of knowledge translation [77], and the other investigated a two-hour online on sleep improvement [78]. Both short online courses were delivered to direct care and support staff from residential, community, respite, acute and primary care settings and changed practices of care (self-reported) and improved perceived quality of care.

#### Summary of higher quality evidence

- In two single group studies, **tele-mentoring** with staff champion support for community-based doctors increased self-reported change in prescribing (level 3), and a programme for health and social care professionals working with people across settings with intellectual disability and dementia improved knowledge (level 2), each evaluated after 12 months of the programmes.
- There was level 2 evidence from a single group study that DEALTS2, a **TTT programme** was noted by trainers to improve empathy in care.
- In small, single group studies evaluating **lecture-based (didactic) models**, an intervention to train HCP in ACPs increased their use (level 3), and a didactic educational workshop increased knowledge of dementia (level 2).

### 3.3 Stakeholder consultation

We consulted eight healthcare professionals from nursing, occupational therapy, general practice, and psychiatry backgrounds, working in primary, secondary and community settings. The group considered that time to train and be trained was the prevailing limiting factor determining how interventions for which we identified evidence of efficacy would work in practice. They noted that mandatory training programmes were already extensive, but rarely related specifically to dementia care strategies that professionals were expected to have learnt during their initial professional training and maintained through self-directed Continuing Professional Development (CPD). They valued group, in-person training as engaging and providing opportunities for sharing experiences. TTT models were valued as enabling tailoring to local contexts.

## Discussion

In our review of English policies on dementia training for healthcare professionals, common themes were a need for role-appropriate training centred around learning objectives defined by the DCSETF, and concerns around patchy implementation of the framework. In our systematic review, priority evidence supported a TTT team-based reflective practice intervention, which improved primary care nurses’ and doctors’ learning (level 2), and self-reported staff practice (level 3) over ≥3 months [22]. Other higher-quality controlled studies which improved outcomes post-intervention supported: a TTT programme for hospital staff which improved client outcomes (agitation) over ≤5 days (level 4) [33]; an expert-led 2-day interactive training for inpatient nurses [37]; and an expert-led, 9-week, occupational therapy-derived training programme for retirement community employees [53]. A controlled study involving three hours of expert-led training for hospital nurses using similar methods did not improve level two outcomes [36]. We consulted eight stakeholders who considered time a limiting factor, but valued group, in-person training to share experiences; and TTT models as enabling tailoring to local contexts.

Successful interventions included lived experience perspectives, experiential learning, case-based and reflective group discussions. These learning strategies reflect professional body recommendations identified in our policy review, for team-based learning, and incorporating lived experience, and the previous review [9]. There were areas where recent evidence extend the recommendations of this previous review. While Surr and colleagues [9,79] advised that successful interventions should involve at least half a day of interaction with a “knowledgeable, skilled and experienced facilitator who is also an experienced clinician or practitioner”, our findings also support TTT models, which can increase reach and enable tailoring and support to local contexts so could support DCSETF implementation.

Interestingly, an intervention for hospital nurses that involved only 3 hours of direct sessions but additional support for implementation from trained staff champions was found to be effective [33], so perhaps TTT models can improve the efficacy of brief training interventions. This reflects findings that, if well supervised and supported, non-clinically trained facilitators can facilitate dementia training to social care workers, where consciously taking a non-expert role can create a positive dynamic of collaborative enquiry [7].

Most of the evidence we found pertained to the training of doctors and nurses. The NHS workforce plan includes a governmental commitment to significantly increase education and training, apprenticeships and alternative routes into professional roles [7]. Staff without professional qualifications comprise just under half of hospital and community healthcare staff [80]; this group may have learning needs that differ in important ways from those who have studied for an obtained a professional qualification, so an absence of evidence around how to support their development of dementia skills is of concern. While team-based learning can be very effective, as discussed in our policy review, the specific needs of diverse professionals must also be met for effective dementia care.

No evidence from high-quality, controlled studies supported self-directed, or didactic training models, echoing Surr et al.’s [79] recommendation to avoid these training modalities. E-learning is a training option that is relatively easy to resource, and its use burgeoned in the pandemic. Perhaps there is scope for development of interactive and effective self-directed formats, which were supported in small, single group studies [57]. Though one study found that in-person learning appeared more lasting than online [24], this is not always practicable and the approach of studies [23,24] that offered a choice of online or in-person formats seems pragmatic.

Effective training is likely to be cost-effective, through enabling people with dementia to communicate their needs, enabling timely diagnosis and management of dementia, comorbidities and care needs, but cost-benefits are yet to be evaluated. Darzi’s report on the NHS emphasised the importance of a well-trained workforce, and transparency of decision-making around expenditure, and that this should include training [8]. Cost-effectiveness studies are needed, to guide service leader and policymaker decisions around funding dedicated training time. Our findings support expert or TTT delivery of interventions that are interactive, rooted in case-based discussions, and preferably delivered to groups in-person. Increasing implementation of such evidence-based training would ensure that people with dementia receive care that is respectful, high quality and equitable.

## Data Availability

No data was produced in the present study

## Acknowledgements

This research is funded through the NIHR Policy Research Unit in Dementia and Neurodegeneration-Queen Mary University of London (reference NIHR206110). The views expressed are those of the author(s) and not necessarily those of the NIHR or the Department of Health and Social Care.

