## Supplementary material for "Dementia Training for Healthcare Professionals: A Systematic Policy and Evidence Review"

Figure 1- PRISMA Flowchart

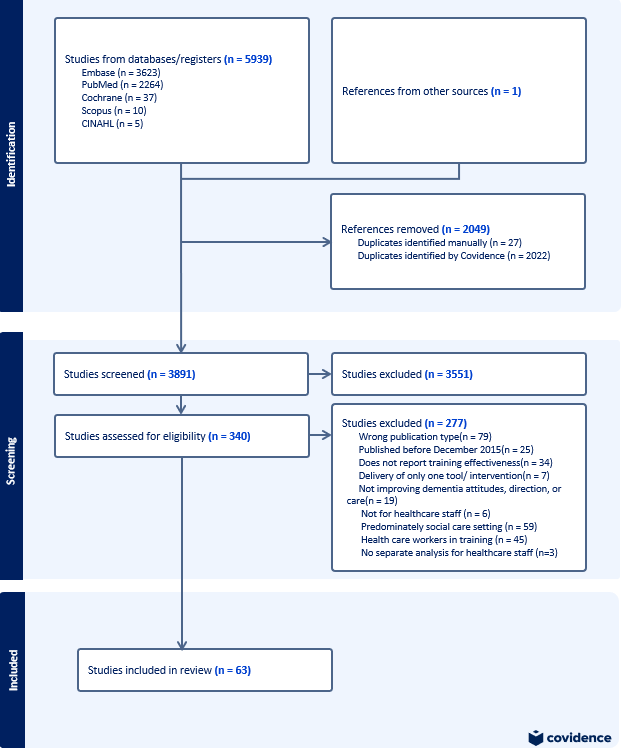

Table S1- Sources search for policy and grey literature

|  | Organisation websites searched |
| --- | --- |
| Governmental and arms length bodies | Department of Health and Social Care (DHSC)  Health Education England  Skills for Care  National Institute for Health and Care Excellence (NICE)  All-Party Parliamentary Group (APPG) on Dementia  NHS England  Care Quality Commission (CQC) |
| Professional bodies | Royal College of General Practitioners  British Geriatric Association  Association of British Neurologists  Academy of Medical Sciences  Royal College of Nursing  Royal College of Occupational Therapists  Royal College of Psychiatrists (RCPsych) |
| Third sector and industry | Alzheimer’s Society  Care Provider Alliance |

Table S2- Search strategy for Pubmed (dates 2015- filter applied)

| Terms |  |
| --- | --- |
| Education | (educat*[Title/Abstract] OR training[Title/Abstract] OR staff development[Title/Abstract] OR professional development[Title/Abstract] OR CPD[Title/Abstract] OR skills training[Title/Abstract] OR curricul*[Title/Abstract] OR learn*[Title/Abstract] OR teach*[Title/Abstract] OR workshop[Title/Abstract] OR module[Title/Abstract]) |

| Staff | ((professional[Title/Abstract] OR staff[Title/Abstract] OR worker[Title/Abstract] OR workforce[Title/Abstract] OR paid carer[Title/Abstract] OR aide[Title/Abstract] OR care worker[Title/Abstract] OR physician[Title/Abstract] OR doctor[Title/Abstract] OR student[Title/Abstract] OR nurse[Title/Abstract] OR therapist[Title/Abstract] OR social worker[Title/Abstract]) |
| --- | --- |
| Dementia | (dementia[Title/Abstract] OR "Frontotemporal dementia"[Title/Abstract] OR korsakof*[Title/Abstract] OR binswanger[Title/Abstract] OR "Progressive Supranuclear palsy"[Title/Abstract] OR alzheimer*[Title/Abstract] OR dement*[Title/Abstract] OR "Korsakoff Syndrome"[Title/Abstract] OR "Wernicke's Encephalopathy"[Title/Abstract] OR "Huntington's Disease"[Title/Abstract] OR "Multi Infarct"[Title/Abstract] OR "lewy bodies"[Title/Abstract] OR "Lewy Body Disease"[Title/Abstract] OR "kluver-bucy syndrome"[Title/Abstract] OR "Vascular dementia"[Title/Abstract] OR "Creutzfeldt-Jakob Syndrome"[Title/Abstract] OR "Alzheimer's Disease"[Title/Abstract])) |

Table S3- Summary of policies and reviews of dementia training for healthcare staff in the UK

| Date | Author | Report Name | Key points relevant to dementia training for healthcare professionals |
| --- | --- | --- | --- |
| February 2015 | Department of Health and Social Care | Prime Minister’s challenge on dementia 2020 [1] | - Expectation that all NHS staff having received training on dementia appropriate to their role. - Adoption of Butterfly scheme in some hospitals which aims at improving people’s experience of staying in hospitals by training staff to offer a positive and appropriate response to people with memory impairment and providing staff with a simple, practical strategy for meeting their needs. - Expectation that all relevant health and care staff who care for people with dementia will be educated about why challenging behaviours can occur and the best way to manage them. |
| 2015 (updated 2018) | Skills for Health, Health Education England and Skills for Care | Dementia Core Skills Education and Training Framework [2] | Benchmarks dementia training for the health and care workforce to support staff to have the right knowledge and skills to deliver high quality, person-centred care for people living with dementia and their carers. It comprises three Tiers, reflecting the role and degree of contact different staff have with people living with dementia. It suggests healthcare staff working regularly with people with dementia and expert leaders undertake Tier 2 or 3. |
| January 2016 | Alzheimer’s Society | Fix dementia care: hospitals [3] | - Dementia awareness and training is essential to enable staff to triage and communicate with people with dementia, their families and carers, understand their needs and offer appropriate care. - For staff who provide direct clinical care, an appropriate level of dementia training is essential to ensure they can understand people’s individual complex needs. - Recommends greater focus on rolling out Tier 2 and 3 training to ensure that NHS staff receive the most advanced support available - Investing in staff training and adaptations to the environment, can save people with dementia the trauma of a fall and the resulting complications. - Recommends all hospitals to publish an annual statement of levels of staff and board dementia awareness and training |
| 2018 | National Collaborating Centre for Mental Health | Dementia Care Pathway [4] | - There is a high variance in staff training, with many feeling unprepared when working with people with dementia and requesting specific training, particularly in working with people with existing comorbidities. - Recommends commissioners to provide referrers (particularly GPs) and providers of support services with education and training programmes - Recommend commissioners to agree training plans and associated costs with providers, engaging local education and training boards as necessary, and ensuring that there are sufficient staff trained to Tier 1, 2 or 3, as appropriate. |
| 2018 | NHS England | Dementia Roadmap [5] | Course developed for primary care staff to help diagnose, support and signpost people with dementia. Offers high quality information about dementia alongside local information about services, support groups and care pathways. |
| July 2018 | National Institute for Care and Excellence | Dementia: assessment, management and support for people living with dementia and their carers (NG 97) [6] | All staff should have training in person-centred and outcome-focused dementia care; with additional face-to-face training and mentoring to staff who deliver care and support to people with dementia, including training on understanding, reacting to and helping people who experience behaviours indicating distress, with follow-up sessions to discuss specific situations, advice on interventions that reduce the need for antipsychotics, and promote freedom of movement. Training should include starting and holding difficult and emotionally challenging conversations. |
| 2018 | The Leeds Beckett University | ‘What works’ in dementia education and training [7] | Review of the evidence shows that the training most likely to be effective has specific features including:   - Tailored and realistic to the role, experience and practice of the learners - Includes specific tools, methods and approaches to underpin care delivery - Presents the experience of living with dementia (video, simulation, etc.) - Is ideally more than half a day duration per subject area (the longer and more in-depth the programmes, positive results more likely) - In case of several sessions, each session should be at least 2hrs - Use of group F2F learning and including interactive learning activities and opportunities for discussion - Avoids didactic methods and self-directed learning - Delivered by an experienced, skilled and flexible facilitator who is also an experienced clinician or practitioner - Delivered in a supportive organisational context, in a dedicated training space where is supportive of good dementia care |
| November 2019 | Royal College of Nursing | Commitment to Care of People living with Dementia: SPACE principles [8] | Dementia training should:   - Focus on values, attitudes and approach of staff, supporting good communication and a relationship-centred approach. - A team approach to training and ensure this is supported in practice by those who have further training. - Include hearing the experience of people with dementia and families/carers. - Be supported by outcomes and data to improve of dementia care rather than blaming individuals - Focus on communication, assessment, reducing risk of developing dementia, life story information, pain, nutrition and hydration, continence, activity, rehabilitation, environment and end of life care. |
| 2020 | National Dementia Action Alliance | Dementia Friendly Hospital Charter [9] | - All staff and volunteers undertake Tier 1 dementia awareness training (within first 3 months of appointment)   All staff working regularly with people with dementia and expert leaders undertake Tier 2 and Tier 3 training appropriate to their role |
| 2021 | NHS England | Dementia wellbeing in the COVID-19 pandemic [10] | Outlined staff training and support needs related to the pandemic, including in the use of remote techniques. |
| February 2022 | Department of Health and Social Care | Dementia: applying All Our Health [11] | Suggests that senior or strategic leads encourage all health and care professionals to complete dementia e-learning or training and becoming a dementia friend. |
| 2022 | NHS England | The Well Pathway for Dementia [12] | The NHS England transformation framework includes “training well”: to develop training for all staff that work with people with dementia in all settings (hospital, General Practice, care home or in the community) and across communities and wider public |
| 2023 | Royal College of Psychiatrists | National Audit of dementia [13] | Reports:   - Large variations of training reported with 0-100% hospital staff with Tier 1 - 58% of hospitals provided figures for staff with Tier 2 training (median 45%)   Recommends: Staff delivering care to people with dementia should have Tier 2 training and it should be recorded. |
| 2024 | NHS England & Alzheimer Europe | Intercultural Dementia Care Guide [14] | Guidance for health and social care workers on delivering culturally appropriate care, signposted to training resources. |
| October 2024 | Care Quality Commission | The state of health care and adult social care in England [15] | - The local dental networks have introduced ‘dementia-friendly’ practices by improving staff understanding of dementia and making simple adjustments to improve the experience for people with dementia and their carers. - Hospital staff do not always understand the specific needs of people with dementia which can negatively affect experience of care. - Training can nurture compassionate and empathetic approach to dementia care. |

Table S4- Characteristics of studies with low quality evidence (MMAT<4)

| **Training for hospital staff** | | | | | | | | | | | | | | |
| --- | --- | --- | --- | --- | --- | --- | --- | --- | --- | --- | --- | --- | --- | --- |
| **Study Country** | **Setting and participants** | **Intervention** | **N** | **Control** | **N** | **Primary outcome** | **Response rate (primary outcome)** | **Outcome timings** | **Outcomes (Kirkpatrick levels)** | | | | **Study design** | **Validity score** |
|  |  |  |  |  |  |  |  |  | **1** | **2** | **3** | **4** |  |  |
| Allegri [16]  Italy | People aged +65 with cognitive function. cared for by hospital staff | IDENTITA: five-hour f2f training based on PCC and delivered by dementia experts using didactic content, slides, assessment tools, and handouts including cases studies and personal experiences | 89 | TAU | 72 | Functional status (MBI) | 42% | BL, PI |  |  |  | 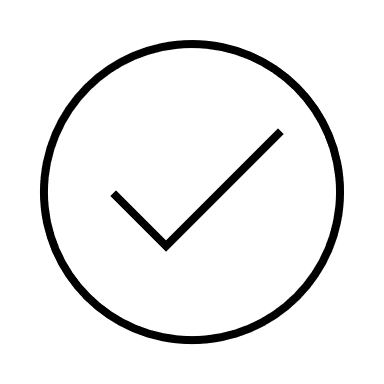 | QN | 3 |
| Gkioka [17]  Greece | Staff members from general hospitals | F2f group two-day (9 hrs) workshop on dementia knowledge | 242 | - | - | No info | 43% | BL, PI, 3 months FU | 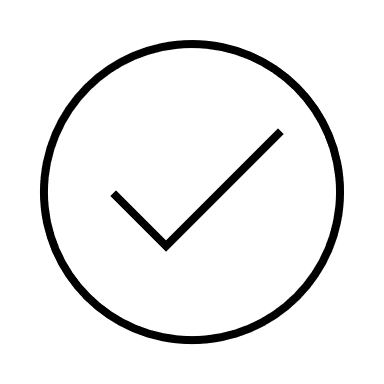 | 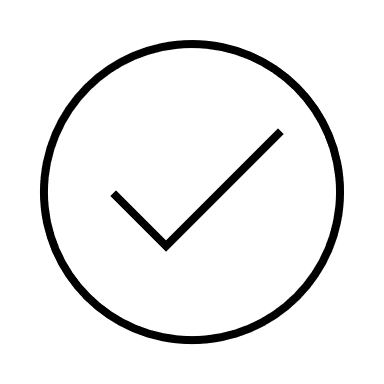 | 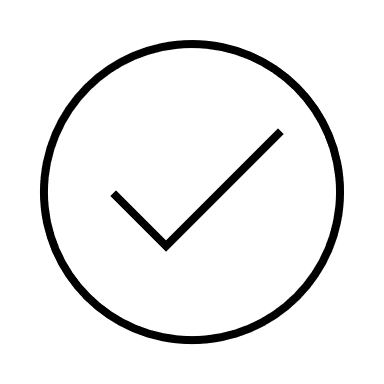 |  | MM | 3 |
| Haffner [18]  USA | Inpatient medical-surgical unit nurses & nurse assistants | Online and f2f nine 20-mintue education sessions focused on dementia types, management of sleep disorders and disturbance in older people | 46 | - | - | Knowledge of sleep disturbances and dementia (measure developed by team) | 59% | BL, PI |  | 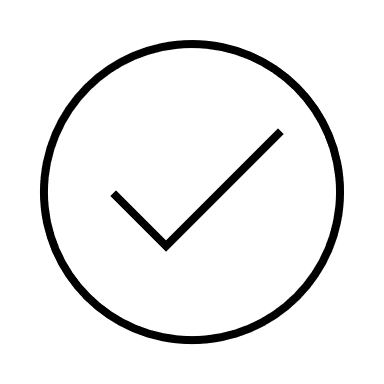 |  | 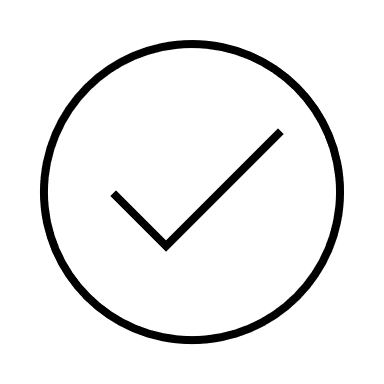^x^ | QN | 3 |
| Jack-Waugh [19]  Scotland | Health and social care professionals employed by an NHS hospital | Dementia champions Programme based on PCC, including pre-reading, five f2f study days; a half day spent in a community setting, distance learning and written assignments; delivered by multi-professional and peer group (including people with dementia) over eight months. | 430 | - | - | No info | No info | BL, PI |  | 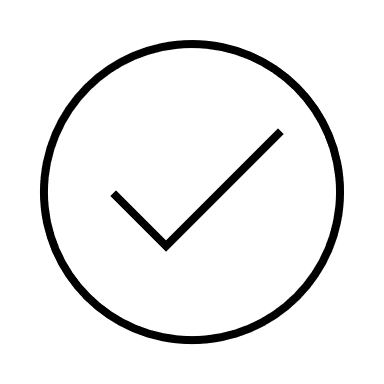 |  |  | QN | 3 |
| La Vallee [20]  USA | Care coordination department staff from a general acute hospital | Referral forms, and educational and local resource information for dementia placed in hospital offices for  two months & sent in sixteen email blasts | 17 | - | - | Knowledge of dementia (DKAS) | 82% | BL, PI | 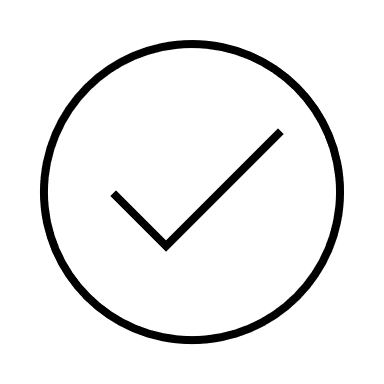 | 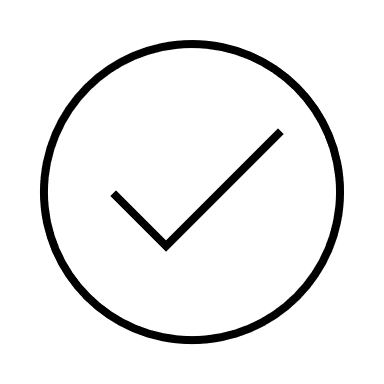 | 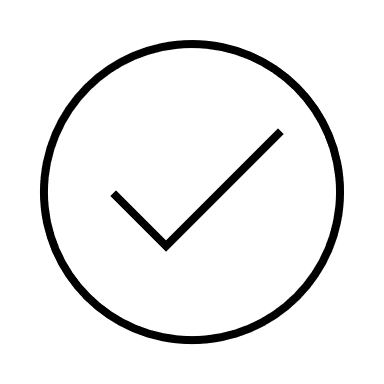 |  | QN | 3 |
| Murray [21]  Australia | Clinical and non-clinical staff from University hospitals | DCHP: dementia education programme (TTT model) delivered by DCHP team over nine months, covering screening for cognitive impairment, use of cognitive impairment identifier above patients’ bedside and communication strategies. | 2587 | - | - | No info | 418/1748 (24%) | BL & 6 months PI |  | 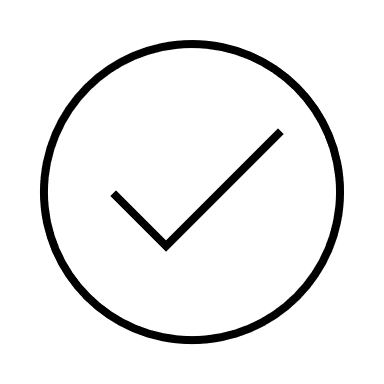 | 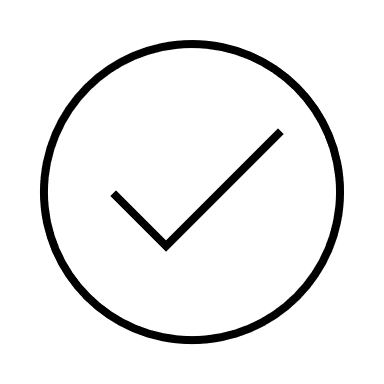 |  | QN | 3 |
| Scerri [22]  Malta | Hospital staff in inpatient rehabilitation setting | Six 90-minute workshops based on an Appreciative Inquiry approach, including PCC and reflective learning about working positively with people with dementia using a story developed from their staff experiences/stories | 68 | - | - | Staff reaction | 57.3% | PI and 4 months FU | 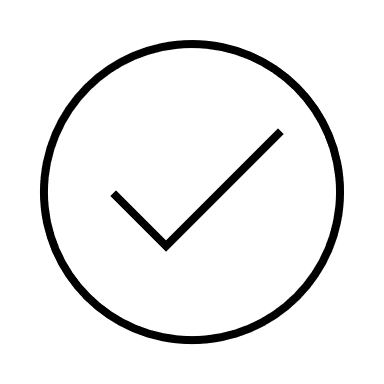 | 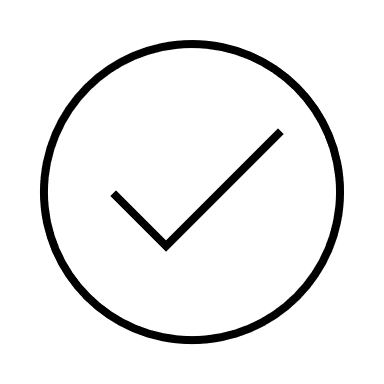 | 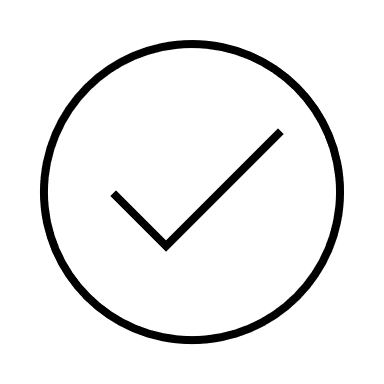 |  | MM | 3 |
| Schindel Martin [23]  Canada | Hospital staff from clinical areas | GPA training: 7.5 hrs of f2f lectures based on PCC principles brain changes in dementia, communication and staff-specific skills delivered by GPA coaches. | 468 | Wait-list | 277 | Self-efficacy in dementia care (SBMSEP) | No info | BL, 8 weeks FU |  | 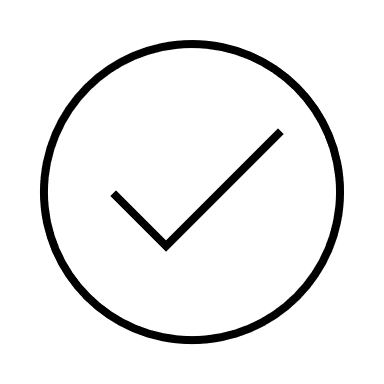 |  |  | MM | 2 |
| Schneider [24]  Germany | Nursing and admin staff from emergency departments | Two-day training covering information on dementia and clinical skills with case studies and group discussions, delivered by the research team | 60 | - | - | Attitude towards (DAS-D) and knowledge of dementia (KIDE) | 34/60 (57%) | BL, 3 & 6 months FU |  | 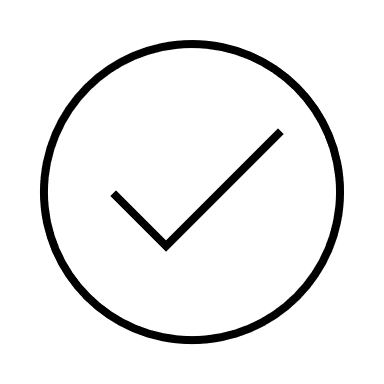 |  |  | QN | 3 |
| Schneider [25]  Germany |  |  | 60 | - | - |  | 9/60 (15%) | BL, 3, 6 & 8 months FU |  | 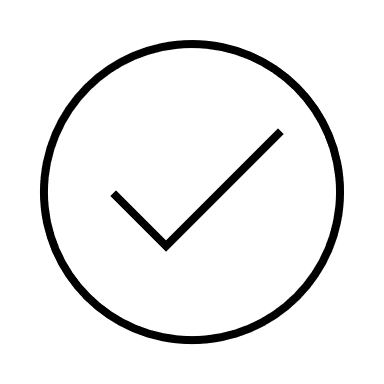 | 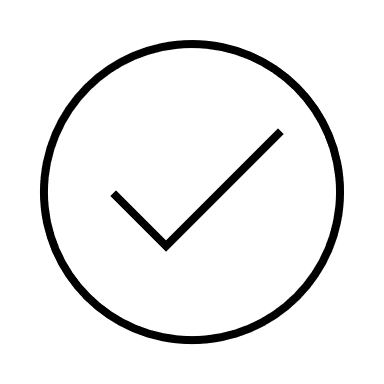 |  | MM | 2 |
| Solecki [26]  USA | Research nurses and nursing assistants from a community-based hospital | VDT: one-hour f2f session including participants wearing sensory devices (Second Wind Dreams) to alter their senses and then given five everyday task to enhance awareness of AD challenges; followed by a 10-15 min debriefing to reflect on the experience; facilitated by nurses/ educators/ care/programme coordinators. | 113 | - | - | No info | 15/113 (13%) | BL, PI, 3-6 week FU |  | 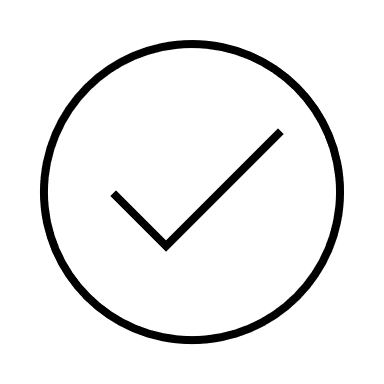 |  |  | MM | 2 |
| Surr [27]  UK | Acute hospital staff | PCTAH: 3.5 hours education by peer facilitators on PCC over 3-4 months; covering types and impact of dementia, emotional, communication, physical health needs, impact of the environment, redefining and supporting challenging behaviours. Delivered in foundation & intermediate levels | 40 | - | - | Staff attitude | 97.5% | BL, 4-6 weeks PI (foundation) and 3-4 months PI (intermediate) |  | 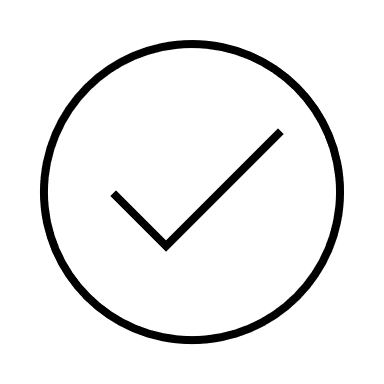 |  |  | QN | 3 |
| Takeuchi [28]  Japan | Dementia care nurses from a psychiatric hospital | Nurse education programme: f2f based on Bandura’s self-efficacy including one-hour lecture on dementia and 3 months of sharing successful experiences based on acquired knowledge and enhancing self-efficacy | 17 | 1: knowledge only  2: TAU | 1:16  2: 18 | Self-efficacy associated with professional knowledge (VAS) & with the programme (GSES) | 51/52 (89.5%) | BL, PI |  | 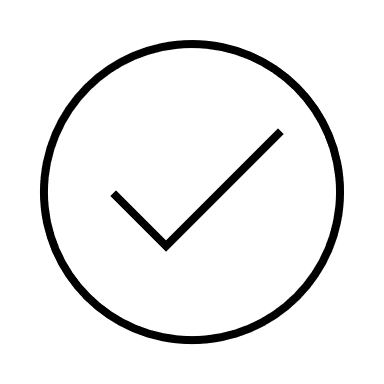 |  |  | MM | 3 |
| Toye [29]  Australia | Nurses, allied health staff, and junior doctors from acute medical ward | Person-CIND: educational modules on dementia in addition to contact, providing resources and pre-planned conversation with patients’ carers over two months. | 59 | - | - | Knowledge of dementia & AD (ADKS, DKAT2) | 44% | BL, PI, 9-12 months FU |  | 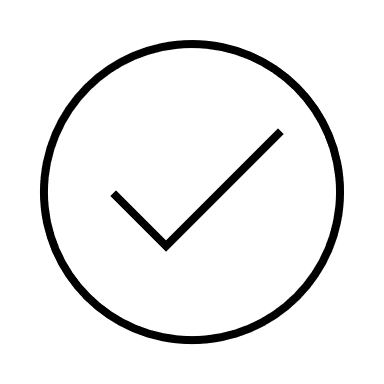 |  |  | MM | 2 |
| **Training for healthcare staff in social care settings** | | | | | | | | | | | | | | |
| Conway & Chenery [30]  Australia | Nurses or nursing assistants in community aged care centres | MESSAGE: one-hour f2f course covering communication strategies in dementia facilitated by a speech pathologist. | 30 | Waitlist | 29 | No info | 64% | BL, PI (active group only), 3 months FU | 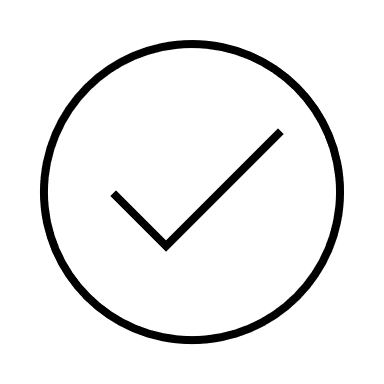 | 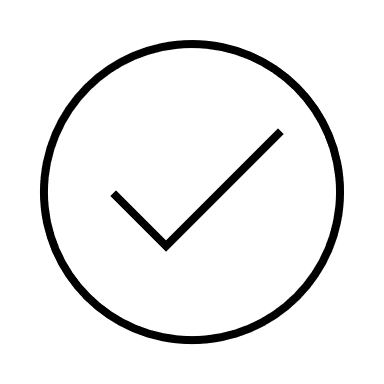 |  |  | RCT | 1 |
| Douglas [31]  USA | Registered Dietitian Nutritionists from community settings | PAC: two-hour f2f education and hands-on practice on sensory abilities, eating and engaging with people with dementia | 25 | - | - | Feasibility & acceptability | 20/25 (80%) | PI | 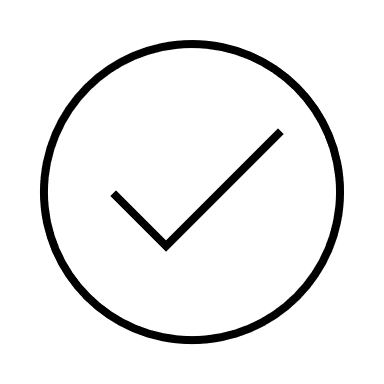 |  |  |  | MM | 3 |
| Duarte [32]  Portugal | Staff of a new community dementia centre | Twelve 45-minute online sessions covering dementia needs and epidemiology, PCC, communication, legal & ethical issues and teamwork delivered by dementia experts | 116 | - | - | No info | 86/116 (85.1%) | BL, PI | 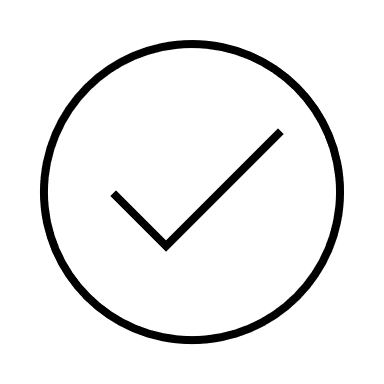 | 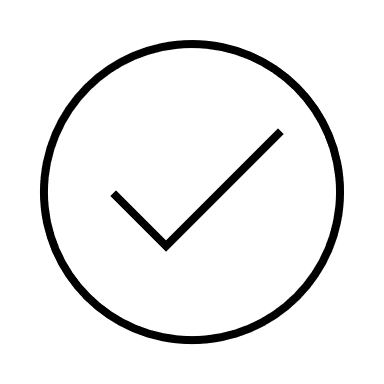 | 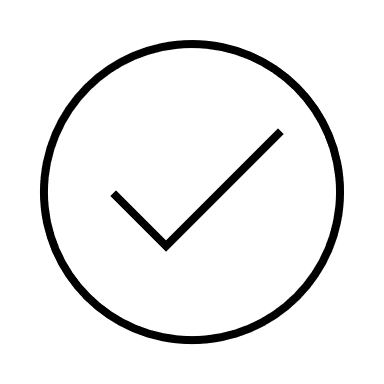 |  | MM | 3 |
| Haydon [33]  Australia | Healthcare professionals in the community | ECHO: tele-mentoring programme eleven monthly one-hour sessions over zoom including a 20-minute didactic presentation, a 30-minute clinical case presentation by a trainee and discussion between the expert panel and learners, concluded with a 10-minute update of the previous month’s case. | 94 | - | - | No info | 44/94 (46.8%) | PI |  | 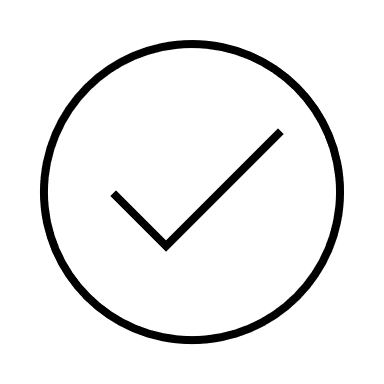 |  |  | MM | 0 |
| Heid [34]  USA | Anyone providing care in community | EFCT: six online lessons (15-30 minutes each) on emotional intelligence, communication and handling one’s and others’ emotions. | 465 | - | - | No info | 53% | BL, PI | 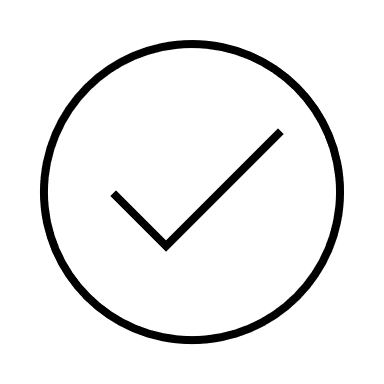 | 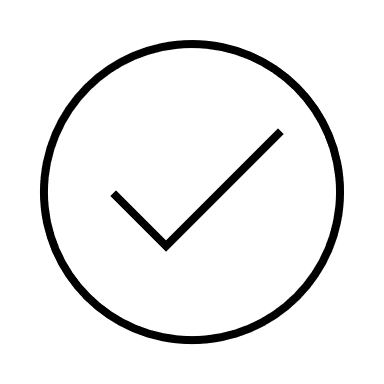 |  |  | QN | 3 |
| Karlin [35]  USA | Care managers and nurses from aging services delivering in-home care | VOICE: two-day f2f training by specialist trainer covering knowledge of dementia, communication, wellbeing, pleasant activities and problem solving, and peer-led biweekly groups, led by service “champions” (who had regular contact with training program) to support implementation of learnings into practice | 14 | - | - | No info | 13/14 (92.9%) | Bl, PI |  | 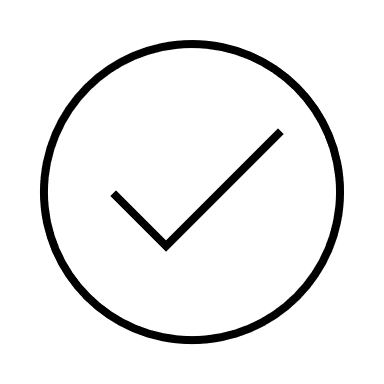 |  |  | QN | 3 |
| Moniz-cook [36]  UK | Community mental health practitioners & champion staff from care homes | Case-specific functional analysis-based training for challenging behaviours: online educational modules on the approach and a decision support e-tool delivered by clinical experts. | 26 | - | - | No info | No info | 6 months | 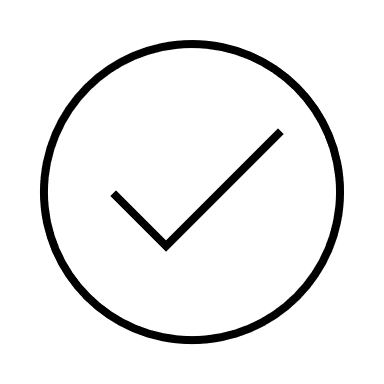 |  |  |  | QL | 2 |
| Nakanishi [37]  Japan | Professional caregivers in long-term care settings | DEMBASE: online 5.5 hours course covering neuropsychiatric symptoms, basic physical needs and environmental sources of discomfort and prescribed medication for nervous system + an interdisciplinary discussion meeting + an online behavioural assessment tool and debriefing | 298 | F2f DEMBASE | 274 | Change in neuropsychiatric symptoms of residents with dementia | 520/572 (91%) | BL, FU |  |  | 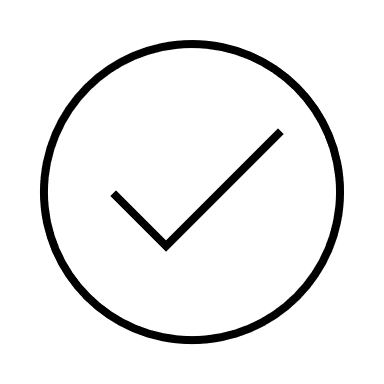 | 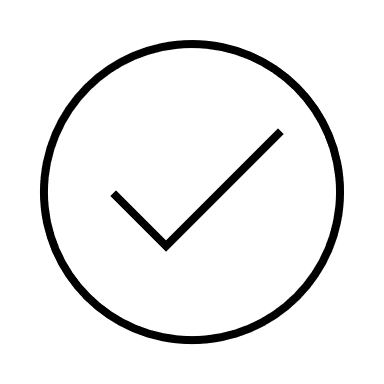 | QN | 3 |
| Plunger [38]  Austria | Pharmacy staff from community pharmacies | Dementia friendly pharmacy: workshop series on communication with people with dementia and caregivers, building networks with local services and pharmaceuticals | 60 | - | - | No info | 41% | BL, PI | 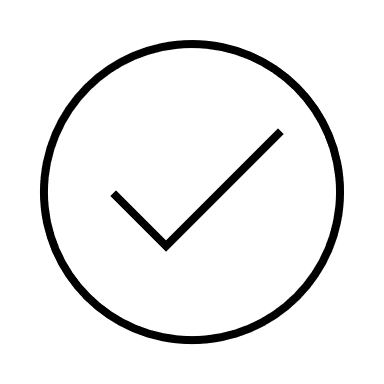 | 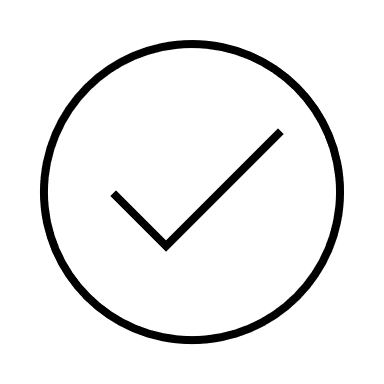 |  |  | MM | 3 |
| Rokstad [39]  Norway | Healthcare and allied staff from nursing homes, special care units and sheltered accommodations, day care centres and home-based nursing care | Dementia ABC educational program: dementia booklet, monthly f2f discussions (90-120 min) and annual workshops covering approaching people with dementia, ethical considerations and engaging with families, and meaningful activities for two years | 1795 | - | - | Competence in PCC dementia (P-CAT) | 580/1795 (32.3%) | BL, mid & PI, 6 months FU |  | 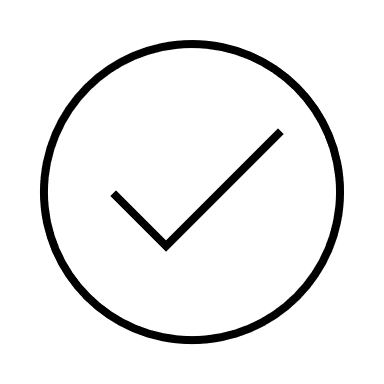 | 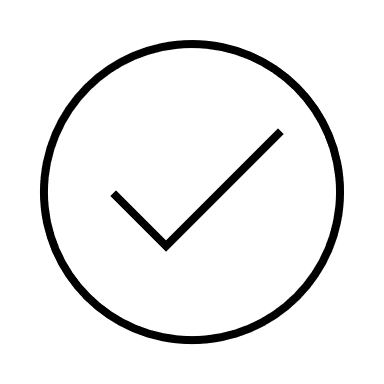 |  | QN | 3 |
| Schneider [40]  USA | Hospice interdisciplinary team members | Aliviado Dementia care- Hospice edition: five online one-hour modules on dementia, depression, pain, management and treatment of BPSD, communication and health care system. | 53 | - | - | Dementia knowledge (DSKA) | 39/53 (73.6%) | BL, PI |  | 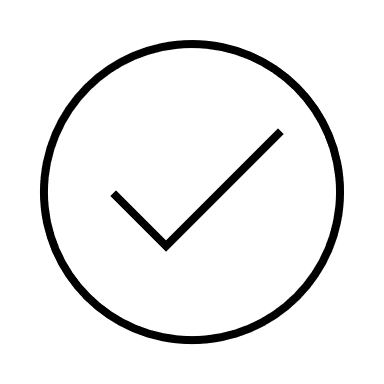 | 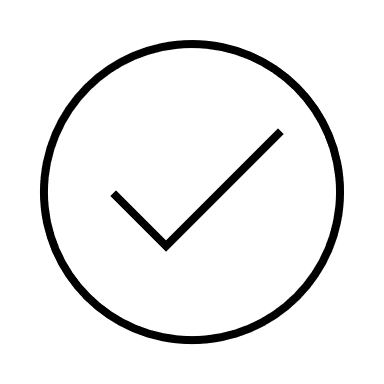 |  | QN | 3 |
|  | **Interventions targeting mixed/secondary care healthcare staff** | | | | | | | | | | | | | |
| Goodenough [41]  Australia | Direct care and support staff from residential, community, respite, acute and primary care settings | Three f2f two-hour workshops on clinical updates in dementia-related behaviours and principles of knowledge translation delivered by the research team | 321 invited | - | - | Change in practice (Conceptual Research Utilization scale and items developed by the team) | 75/321 (23.3%) | 6 months PI |  |  | 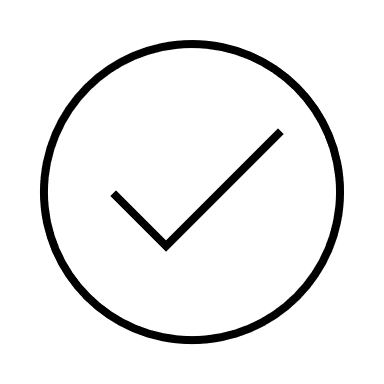 |  | QN | 3 |
| Goodenough [42]  Australia | Direct care and support staff from residential, community, respite, acute and primary care settings | Online two-hour course over two weeks covering introduction and strategies and tips for improve sleep for both residents and staff | 1186 invited | - | - | Change in quality of care (PIQOC) | 161/1186 (13.5%) | 3 months PI |  |  | 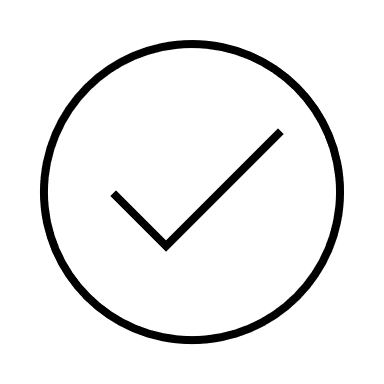 |  | MM | 3 |
|  | **Trainings for primary care staff** | | | | | | | | | | | | | |
| Jennings [43]  Ireland | Primary care professionals | Three-hour f2f interactive workshop covering professionals roles and responsibilities after a dementia diagnosis, team collaboration, information on diagnosis & inter-professional communication skills | 54 | - | - | No info | No info | BL, PI | 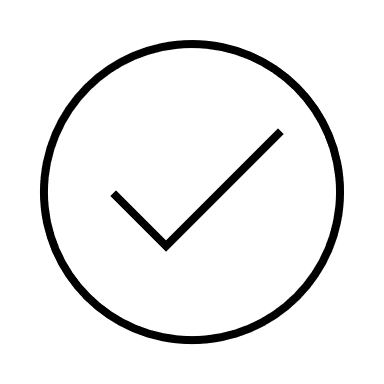 | 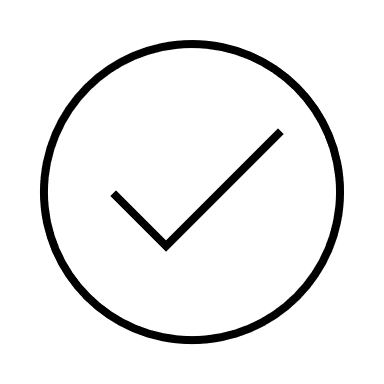 | 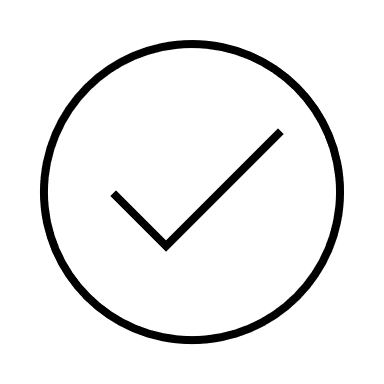 |  | MM | 2 |
| Kistler [44]  USA | Primary care clinicians | Four-hour f2f training on ACP in dementia, end of life, decision-making | 51 | - | - | No info | 33/51 (88%) | BL, PI | 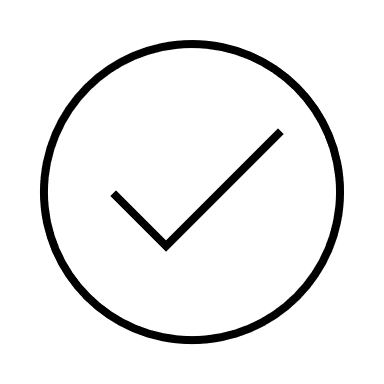 |  | 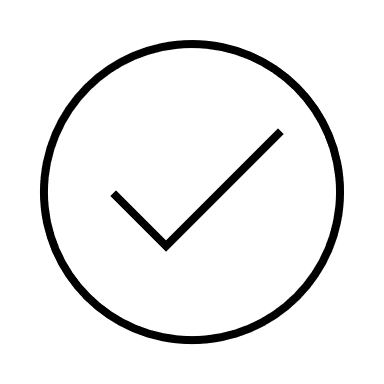 | 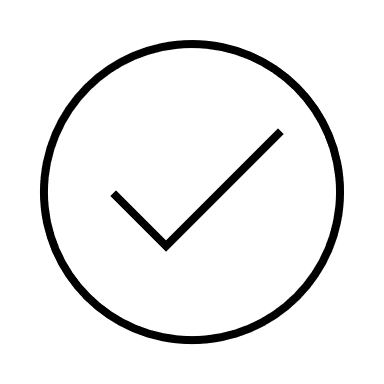 | MM | 3 |
| Kosteniuk [45]  Canada | Primary care staff | Continuing Education: three webinars (60-90 minutes) on medication or drug-induced cognitive impairment, management of challenging behaviour and legal capacity using presentations and discussions | 68 | - | - | No info | 46/68 (67.6%) | PI | 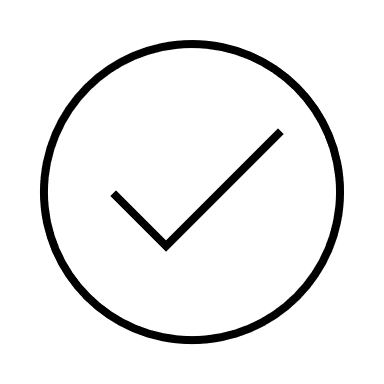 |  |  |  | MM | 3 |
| Lee [46]  Canada | Family medicine residents | Dementia care training delivered by family physicians instead of geriatrics experts; training for faculty to best support residents, three-hour online tutorial on assessment, diagnosis and management + experiential learning opportunities. | 98 | TAU | 35 | No info | No info | PI | 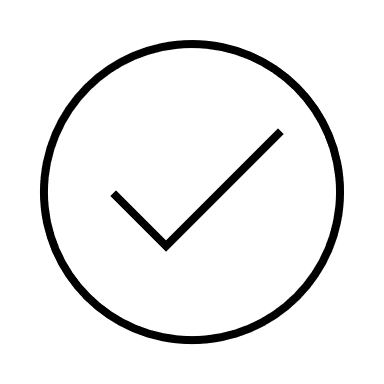 | 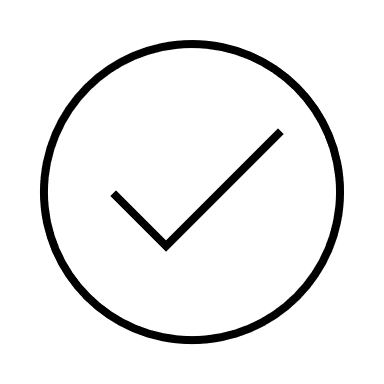 |  |  | QN | 3 |
| Mellinger [47]  USA | NCMs in Primary Care settings | CRESCENT: Six online one-hour modules covering medication, behaviour management, safety assessment, carer wellbeing, community resources, decision making and advance care planning, and one day of live virtual learning delivered by a geriatrician and a geriatric psychiatrist with ongoing support | 11 NCM, 26 person with dementia | No CRESCENT | 12 NCM, 28 person with dementia | Uptake (Documentation of CRESCENT protocols) | No info | 6 months PI |  |  | 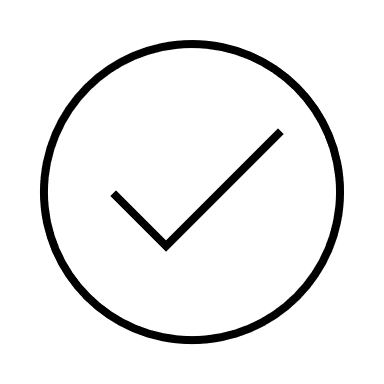 |  | RCT | 2 |
| Ollerenshaw  [48]  Australia | GPs and practice nurses | DTP: online decision tree and associated information to assist with diagnosis, referral and care planning for dementia | 42 | - | - | Awareness and usage of the tool | 42/423 (10%) | 18 months FU | 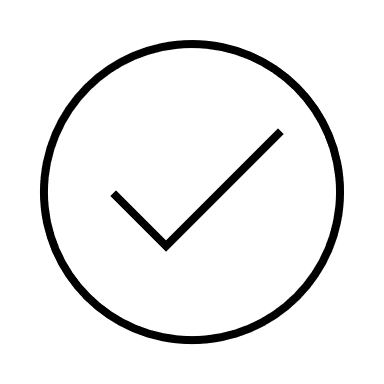 | 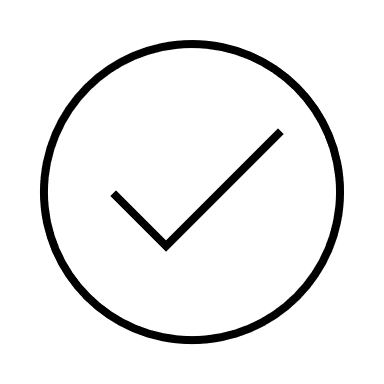 |  |  | MM | 2 |
| Sass [49]  UK | Primary care staff | PG-Cert online and f2f two twelve-week modules on dementia assessment, diagnosis and intervention with mentoring by a memory-service assessment staff | 24 | - | - | No info | No info | PI | 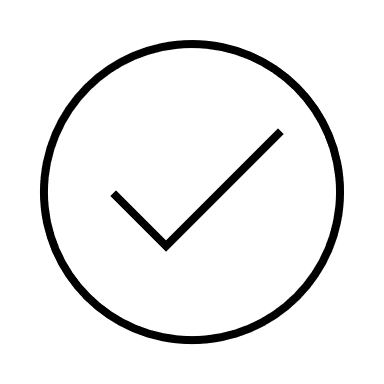 | 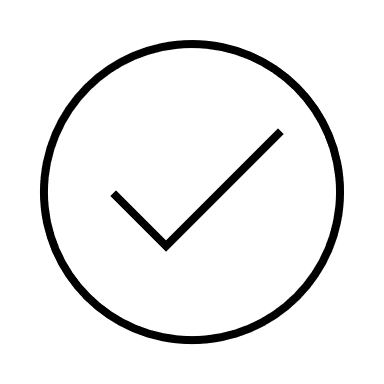 | 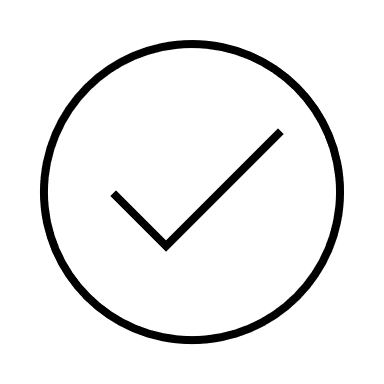 |  | MM | 1 |
| Wang [50]  China | Primary care nurses | F2f TTT model 20-hour weekly training covering diagnosis, PCC, supporting families, comorbidities, behaviour management, safety, treatment and knowledge translation using lectures, case studies, group discussions and presentations delivered by the project team | 90 | Waitlist | 92 | No info | 170/182 (93.4%) | BI, PI & 3 month FU | 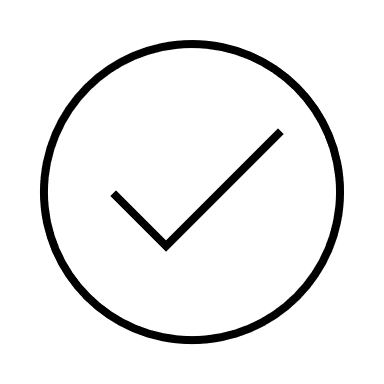 | 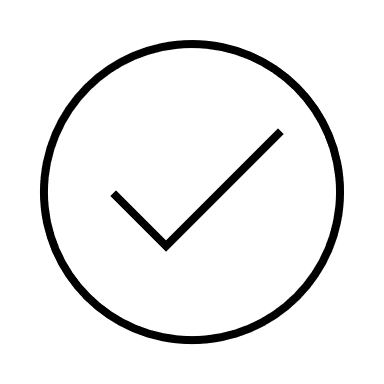 | 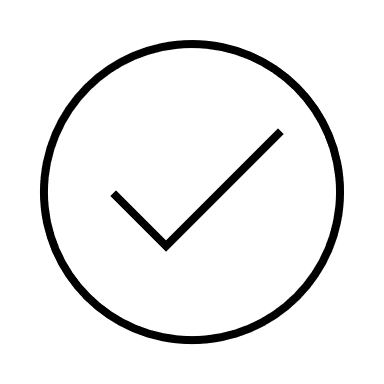 |  | RCT | 3 |

**Legend:** IDENTITA- Italian Dementia-Friendly Hospital Trial; f2f- Face to face; PCC- Person-centred care; TAU- treatment as usual; MBI- Modified Barthel Index; BL- Baseline; PI- post-intervention; QN- Quantitative; FU- Follow-up; MM-Mixed-methods; x: changes in patients’ sleep patterns was an outcome. However, the small sample did not allow statistical analysis; DKAS- Dementia Knowledge Assessment Scale; DCHP- Dementia Care in Hospitals Program; TTT- Train the trainer; GPA- Gentle Persuasive Approaches; SBMSEP- Self-Perceived Behavioural Management Self-Efficacy Profile; DAS-D- Dementia Attitudes Scale; KIDE- Knowledge in Dementia; Virtual Dementia Tour; AD- Alzheimer’s Disease; PCTAH- The person-centred care training programme for acute hospitals; VAS- Visual analog scale; GSES- Generalised self-efficacy scale; Person-CIND- Person-Focussed and Caregiver-Informed Nurse-Driven model; ADKS- Alzheimer's Disease Knowledge Scale; DKAT2- Dementia Knowledge Assessment Tool Version 2; MESSAGE- Communication strategies in dementia for care staff training programme; RCT- Randomised Controlled Trial; PAC- Positive Approach to Care; ECHO- Extension for Community Healthcare Outcomes; EFCT- Emotion-Focused Communication Training; VOICE- Vital Outcomes Inspired by Caregiver Engagement; QL- qualitative; DEMBASE- psychosocial Dementia Behaviour Analysis and Support Enhancement; P-CAT- person-centred care assessment tool; BPSD- Behavioural and Psychological Symptoms of Dementia; DSKA- Dementia Symptom Knowledge Assessment; PIQOC- Perceived Improvement in the Quality of Care; ACP- Advanced Care Planning; NCM- Nursing Care Managers; CRESCENT- CaReEcoSystem primary Care Embedded dementia Treatment; GP- General Practitioner; DPT- Dementia Pathway Tool; Postgraduate certificate.

[9] National Dementia Action Alliance. Dementia Friendly Hospital Charter. Published online 2018.

[10] NHS England. Dementia wellbeing in the COVID pandemic. Published online 2021.
